# The Risk of Lifting COVID-19 Confinement in Mexico

**DOI:** 10.1101/2020.05.28.20115063

**Authors:** Cristy Leonor Azanza Ricardo, Esteban A. Hernandez-Vargas

## Abstract

The novel coronavirus SARS-CoV-2 has paralysed our societies, leading to self isolation and quarantine for several days. As the 10th most populated country in the world, Mexico is on a major threat by COVID-19 due to the limitations of intensive care capacities, and a total of about 1.5 hospital beds for every 1000 citizens. In this paper, we projected different scenarios to evaluate sharp or gradual quarantine lifting strategies, however, even in the hypothetical scenario that Mexico would continue with full confinement, hospitals would be reaching the maximum capacity of hospital bed occupancy. Mexican government is planning to relax the strict social distancing regulations on 1 June 2020, however, epidemic rebound risks are latent.

Our results suggest that lifting social confinement needs to be gradually sparse while maintaining a decentralized region strategy among the Mexican states. To substantially lower the number of infections, predictions highlight that the elderly should remain in social confinement (approximately 11.3% of the population); the confined working class (roughly 27% of the population) must gradually return in at least four parts in consecutive months; and to the last the return of students to schools (about 21.7%). As the epidemic progresses, de-confinement strategies need to be continuously re-adjusting with the new pandemic data. Assuming the most optimistic scenario by our predictions, the smallest number of new COVID-19 cases, Mexico would require at least a 3 fold increase in hospital capacities dedicated for COVID-19. Furthermore, to observe the real dimension of the epidemic, Mexico would need to increase to at least 18 samples per 1000 people, currently is only 0.6 per 1000.

All mathematical models, including ours, are only a possibility of many of the future, however, the different scenarios that were developed here highlight that a gradual decentralized region de-confinement with a significant increase in healthcare capacities is paramount to avoid a high death toll in Mexico.

## Introduction

Severe Acute Respiratory Syndrome Coronavirus 2 (SARS-CoV-2) is the virus behind the 2019 coronavirus disease (COVID-19) with alarming levels of spread and death tolls worldwide. With more than 4 millions confirmed cases and 292,046 deaths^1^, COVID-19 pandemic has spread 212 countries moving the epicentre from China to Europe and consequently to America^1^. While potential vaccines and antiviral drugs are under fast development^2^, epidemiological models have underlined the relevance of social distancing interventions as the main weapon so far to mitigate COVID-19 pandemic^3^.

Epidemiological models have played a central role to advance our understanding in SARS-CoV-2 transmission^3–6^. In its early stages, the epidemic can double in size every 7.4 days^7^. The case fatality rate for COVID-19 ranges from 0.3–1%^1^ up to 20%^8^. The basic reproductive number has been computed roughly 2.2 (95% CI, 1.4 to 3.9)^4,7^. The lesson learned from China, Italy, and the United States pointed out that COVID-19 can quickly result in high demands of healthcare capacities deriving in the collapsing of hospitals of well-resourced nations^9,10^.

Developing countries with limitations of intensive care beds are highly jeopardized by COVID-19. Mexico ranks as the 10th most populated country in the world with about 127 million people^11^. About 60.4% of the population is economically active whereof 56.2% depends on the informal workforce^11^. Albeit the economic impact and levels of moderate poverty, Mexico adopted on 15 March 2020 social confinement, which was relatively early compared to the confirmed number of cases and deaths^12^. By the cut-off date of 25 May 2020, the public health strategy in Mexico has resulted in 71,105 infected cases and 7,633 deaths by COVID-19^12^.

The public health strategy followed by Mexico for the COVID-19 pandemic is the sentinel model, which was implemented by Mexico in 2009 against the pandemic of influenza H1N1^13^. The sentinel model consists on three phases. In stage 1 of the outbreak the model applies case-by-case monitoring. At phase 2, the model focuses on places where the epidemic is growing, at this stage a community-based surveillance is applied. In Phase 3, which is the peak infection phase, the central problem of pandemic surveillance is not to monitor the growth of the pandemic but to ensure that hospital capacities are not exceeded. Mexico has an extremely limited healthcare capacity, approximately 1.5 bed for every 1000 citizens. Based on an early data of the pandemic and assuming social confinement restrictions, the Sub-secretary of Prevention and Health Promotion of Mexico presented mathematical model predictions with a peak of the pandemic Mexico between 8–10 May and an speculative end of the pandemic on 25 June 2020^12^. Mexico is planning to lift the strict social distancing regulations on 1 June 2020. However, an abruptly lifting of social confinement would likely result in new waves of new COVID-19 cases and high death tolls.

In this work, we fit a derivation of the SEIR model for COVID-19^14^ using data of the COVID-19 epidemic in Mexico^12^ as well as its public health capacities and demographic conditions^11^. Different lifting confinement scenarios are evaluated for the main regions of Mexico in order to inform public makers to tailor decentralized region strategies through the Mexican territory with the ultimate goal to minimize deaths.

## The Sentinel Model for Tracking COVID-19 in Mexico

A generalized SEIR model similar to Peng *et al*.^14^ is developed to tailor lifting social confinement scenarios to different regions of Mexico. Different assumptions need to be integrated in order to match the sentinel model used by Mexico against COVID-19^13^. The model writes as follow:

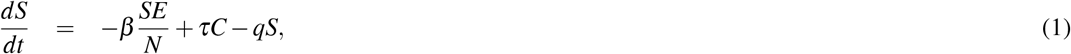

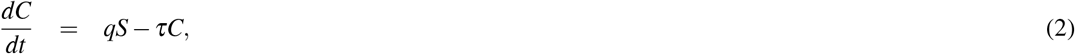

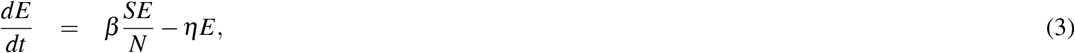

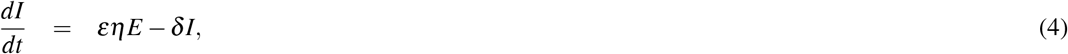

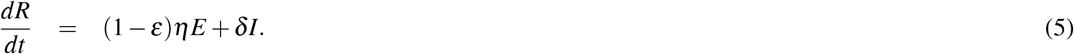

The susceptible population is represented by *S*, from which the total country population (*N*) is about 127 million^11^. The exposed population to the virus is considered by *E*. In the sentinel model applied in Mexico, testing is about 10% of the suspected cases while 100% of hospitalized cases and deaths. The infected cases (*I*) once reported by the Mexican Government are in a hospital or at home under confinement. Therefore, it is a reasonable assumption to consider that new infections are mainly driven by the exposed population with the term 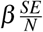. The asymptomatic phase has been reported on average 5–6 days, but it can be up to 14 days^1,3^. The exposed population will leave this compartment with a rate *η*. A fraction *ε* of the exposed population that will present severe symptoms or complications, and consequently counted in the reports of infected cases (*I*). This is approximately 20%^1,3,15^, that is *ε* = 0.2. The other fraction 1 *− ε* will move to the compartment of remove (*R*), those cases that recover or die. The average infection time is about 3–7 days^16^, while the clinical recovery is 2–6 weeks after symptoms^17,18^. Thus, we assume the recovery rate of infected cases (*δ*) in 10 days.

The first confirmed case of COVID-19 in Mexico was reported on February 28th, 2020^12^. At the day of social confinement, 15 March 2020, the number of infected cases (*I*_0_) for the whole country were 12^12^. Due to the change in social movement as well as the very low number of infected cases between the first confirmed case till the day of social confinement, we perform our parameter fitting at the first day of social confinement. The initial number of exposed population (*E*_0_) is approximately a factor *f* of 8 to 12 respect to the confirmed cases^19^, that approximates *E*_0_ = *fI*_0_.

The compartment *C* is the confined population which entered with a rate *q* from the susceptible population. *τ* the effect of population de-confinement, which can be attributed to government policies as well as resistant/forgetting factor by the population to keep adequately the confinement. Estimations by the COVID-19 community mobility report of Google^20^ suggest a reduction in the mobility of about 50 to 70%. Thus, the ratio between *τ/q* = 2*/*3 would represent the 60% confinement percent in the steady state. If we consider a fast confinement rate, then *q* = 1.

A re-sampling strategy was employed to fit the parameters *β* and *η*. The range of parameter values in Table 1 served to generate 3000 sets of random parameters with specific statistical distributions as presented in Figure A.1. This set of parameters consequently was used to fix parameters and perform 3000 fitting repetitions, which minimize the root mean square (RMS) difference between the model predictive output 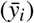, and the reported data of infected cases (*y_i_*). Among different several stochastic optimization algorithms, the minimization of RMS is performed using the Differential Evolution (DE) algorithm^21^ with our Python library *PDEparams*^22^. Using the python library *http://st.t.interval* and parameter fitting distributions based on 3000 samples, the 95% confidence interval (CI) is computed. The embedded panel in Fig. 1 present the best fit and sampling procedure for the whole country, Mexico City (CMX) and the State of Mexico (MEX), while the other regions with major number of infected cases is shown in Fig. A.2.

**Table 1.** Parameters of the Sentinel Model (1)-(5) for Tracking COVID-19 in Mexico. Parameters.

| Parameters | Value (Range) | Reference |
| --- | --- | --- |
| Exposed population with severe symptoms, $\varepsilon$ | 0.2 (0.15-0.3) | 1, 3, 15 |
| Recovery time, $\delta$ | 2 (1.5–3) weeks | 17, 18 |
| Factor for estimate exposed cases, $f$ | 10 (8 to 12) | 19 |
| Reduction in the mobility $C_S$ | 60 (50-70)% | 20 |
| Confinement rate, $q$ | 1 (0.9-1.1) | Fixed |
| Infection rate, $\beta$ | 1.25 | Estimated |
| Incubation period, $\eta$ | 0.45 | Estimated |
| Reproductive Number, $R$ | 2.8 (2.2-3.2) | Estimated |

**Figure 1.**
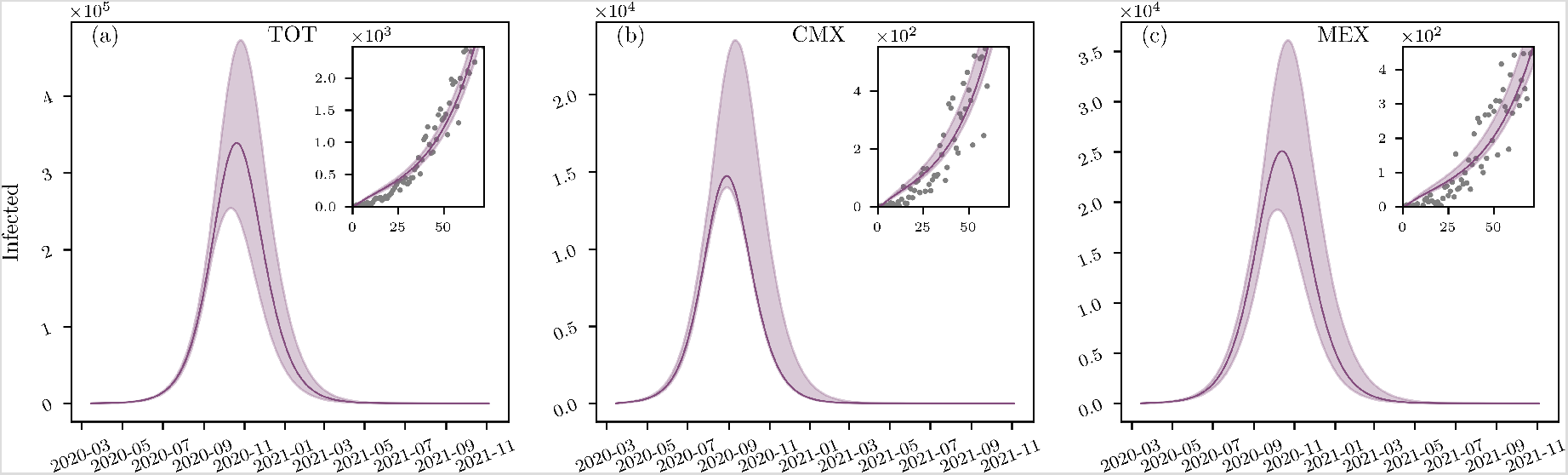
SEIR model (1)-(5) fit and and its respective prediction for the country, CMX and MEX. Model fitting is performed at the initial day of social confinement 15 March 2020). Shadows indicate the range of 3000 fits of infected cases considering the range of parameters in Table 1.

Pandemic predictions underlined that while the early social confinement would delay the initial expected peak of the pandemic on middle may^12^ till July-August, the number of infected per day would reach very high numbers, order of 10^4^-10^5^. In fact, reproductive numbers for the whole country as well as major regions affected with COVID-19 were between 2.3 and 3.6 (see Table A.1). This underlines that even social restrictions SARS-CoV-2 was still spreading among the country and major populated cities. The heterogeneity of the infections cases and its respective public health necessities varied significantly among different geographic regions of Mexico. Considering the reported number of infected cases, Figure 2 presents the percent of the COVID-19 infected cases that require Intensive Care Units beds (ICU), intubation facilities (INT) and hospital beds (HOS) from 12 April till 25 May 2020^23^. Large box plots imply that the requirements of the state was changing a lot in time *e.g* Zacatecas (Zac) and Colima (Col). On the other hand, small box plots highlight that public health requirements for the state were very consistent during the reported period *e.g* CMX.

Assuming the percents of required hospital beds respect to the reported infected cases are consistent in the future (about 38% for the whole country, see Figure 2 and Table A.2), the most optimistic peak of the pandemic presented in Figure 1 could be about 254,174 infected cases, which would result in a saturation of hospitals. That is, the whole country would require to have about 97,180 hospital beds, while the number available of beds for COVID-19 pandemic reported for the whole country is about 49,083^24^. This is a major public concern as Mexico would only be able to provide approximately 50% for the peak of COVID-19 pandemic, otherwise hospitals capacities for non-COVID-19 patients would be reduced.

**Figure 2.**
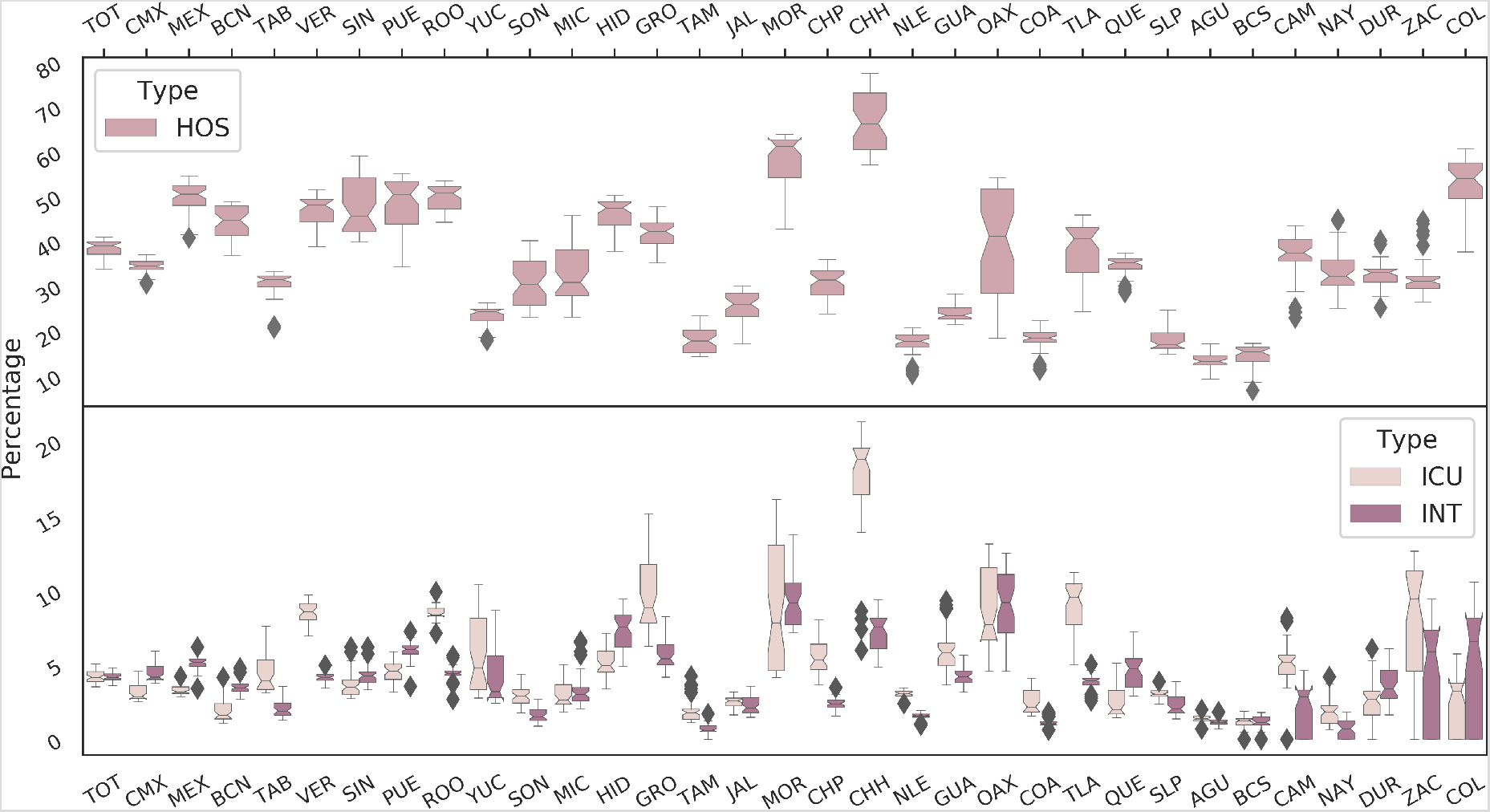
National requirements of Intensive Care Units beds (ICU), intubation facilities (INT) and hospital beds (HOS). These percents are based on the reported data of infected cases that require ICU, INT or HOS. Box plots are composed by data sets from 12 April till 25 May. Therefore, those states with small box plots reveal a very stable needs in public health capacities in time (*e.g* CMX). Data was extracted from^23^.

## Strategies for Social Confinement Lifting

Mexico is planning to lift social distancing restrictions on 1 June 2020^12^. Based on the nominal values presented in Table 1, we develop different possible scenarios to gradually lift social confinement. Panels (a), (b) and (c) in Figure 3 present a simulation scenario with a sharp and massive return of activities on 1 June 2020. This would result in a major increase of infected cases, about two order of magnitude higher than the scenario with social confinement in Figure 1.

**Figure 3.**
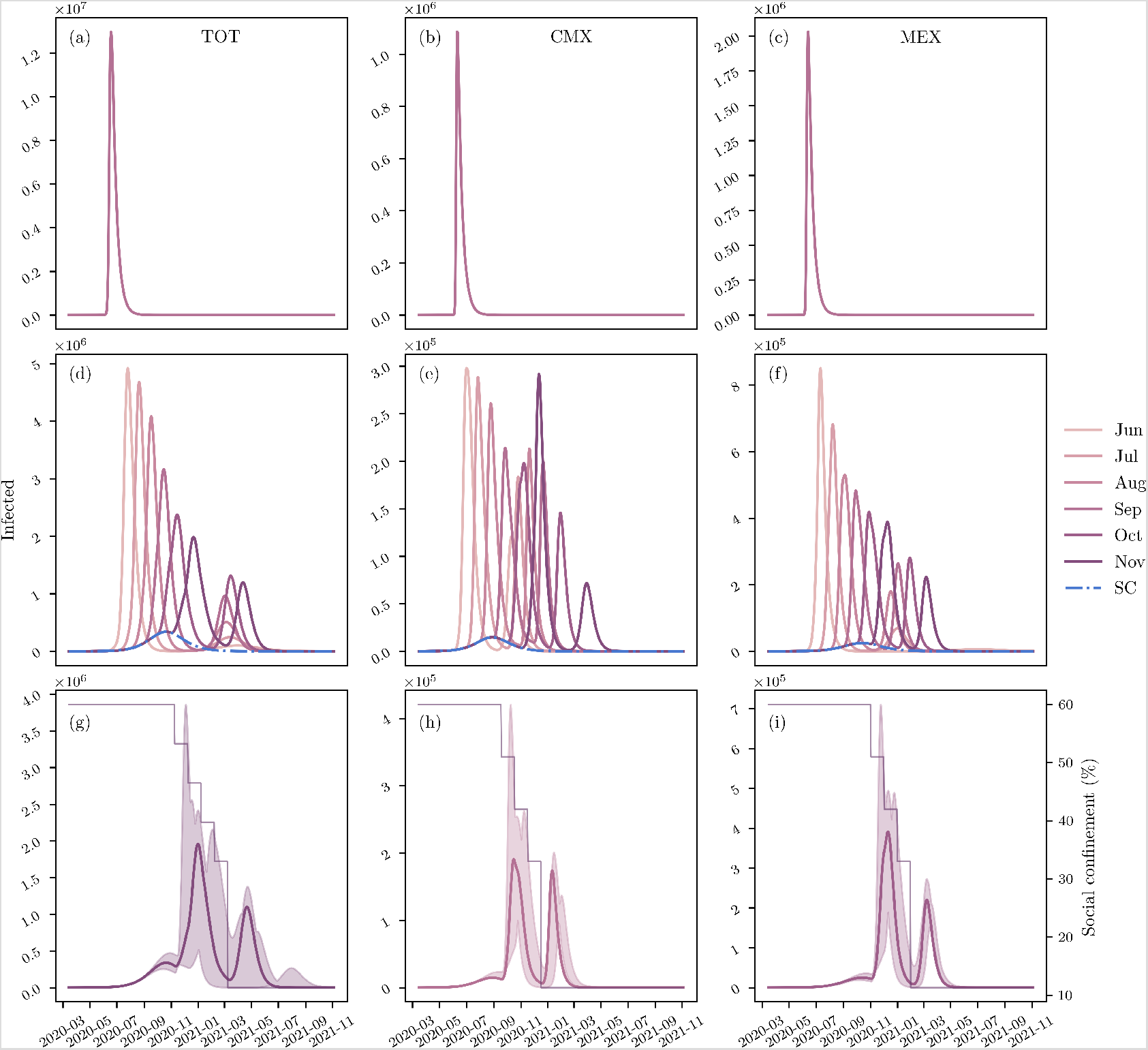
Confinement Lifting Strategies. Panels (a), (b) and (c) present a sharp and massive return of people for social de-confinement on 1 June 2020 for the whole country, CMX, and MEX, respectively. Panels (d), (e) and (f) shows a step-wise social confinement lifting at different time points. This consists on keeping the elderly in confinement, while the confined working force of Mexico (about 27%) returns to activity in four parts (three steps for CMX and MEX), each one in consecutive months, while the students would come back to school one month after the working group. Panels (g), (h) and (i) underlined the best scenario for confinement lifting in the whole country, CMX, and MEX, respectively.

Considering that the confinement in Mexico was about 60% of the population^20^, we evaluate a step-wise social confinement lifting based on a gradual de-confinementof different age groups. Based on Mexican reports by the INEGI^11^, Mexican population can be divided into three major age groups: *G*_1_ are those between 0–24 years old (approximately 42.94% of the population), *G*_2_ are those between 25–59 years old (approximately 45.75% of the population), and *G*_3_ are those between 60- years old (approximately 11.32% of the population). Thus, we consider that the elderly, group *G*_3_, should stay in social confinement. The remaining population in confinement, about 48.7%, is divided into two groups: the first are students from 3 to 17 years old composed approximately 21.7%^11^, the second is the group of the working force of Mexico that stayed in social confinement, around 27%^11^.

The initiation of activities of the working class and students is tailored to reduce the peak of the pandemic. Figure 3(d,e,f) shows a step-wise de-confinement strategy starting at different months (from June till November), returning the working class in four consecutive months (or three for CMX and MEX) and at the last the returning of students to schools. Independently of the initiation time of de-confinement, predictions in Figure 3 highlight that confinement lifting must be avoided before passing the peak of the pandemic. If the initiation of activities is before the first peak, this would result in a much higher number of infected cases, and consequently death tolls. Similar outcomes are obtained for CMX and MEX (Figure 3(e,f)) as well as in major regions of Mexico (Figure A.3). Figure 3(d,e,f) underlines that the best time to initiate de-confinement in CMX and MEX would be between October and November. While this step-wise confinement lifting strategy is the best alternative to mitigate the peak of the pandemic, it would still derive a second wave of the pandemic equivalent to the first one under social restrictions. Estimations of Hospital requirements for de-confinement strategies for different Mexican regions are presented in Table A.2.

## Discussion

The year 2020 has revealed one of the biggest pandemics reported in history, the novel coronavirus SARS-CoV-2 causes a severe and potentially fatal acute respiratory syndrome (COVID-19). Epidemiological reports by countries with strong public health capacities have uncovered that potential of COVID-19 to saturate hospitals in short time^1^. Therefore, COVID-19 is a major threat to developing countries because of limitations of intensive care beds. While no vaccine or antiviral drug is likely to be available soon, the only remaining tool against COVID-19 is social confinement. However, a prolonged lock-down will hugely affect societies, education, and economy.

In this paper, we take as a case study Mexico, which adopted on March 15 social confinement to avoid the fast spread of the virus and the eventual collapse of the public health services^12^. Mexico is a highly populated country with major levels of moderate poverty. A strict social confinement has been extremely difficult to apply in a country with 56.2% of the population working informally^11^. Mexico will lift the social confinement on 1 June 2020, however, the consequent waves of infections are eminent and an abruptly de-confinement would result in high death tolls in a limited public health system. On March 24, an official communication^24^ reported that 40% of the available hospital beds in the country were dedicated to the COVID-19 epidemy. That is, a total of 49,083 hospital beds, 256 intensive care units, 5,523 ventilators and 2,446 intensive care beds. Furthermore, a re-conversion program is in process, to increase the health capacity^24^.

In spite of social confinement, our results point out a high threat to Mexico (Figure 1), that is for the most optimistic scenario, the pandemic peak would be about 338 thousand infected people. If the ratios of hospitalization observed so far are consistent in the next months (about 38% of infected cases, see Figure 2), Mexico would be almost at the maximum capacity, approximately 98% hospital bed occupancy, despite the fact that beds of non-COVID-19 patients are considered for COVID-19 patients. Therefore, social confinement lifting would very likely result in a hospital system breakdown.

**Figure 4.**
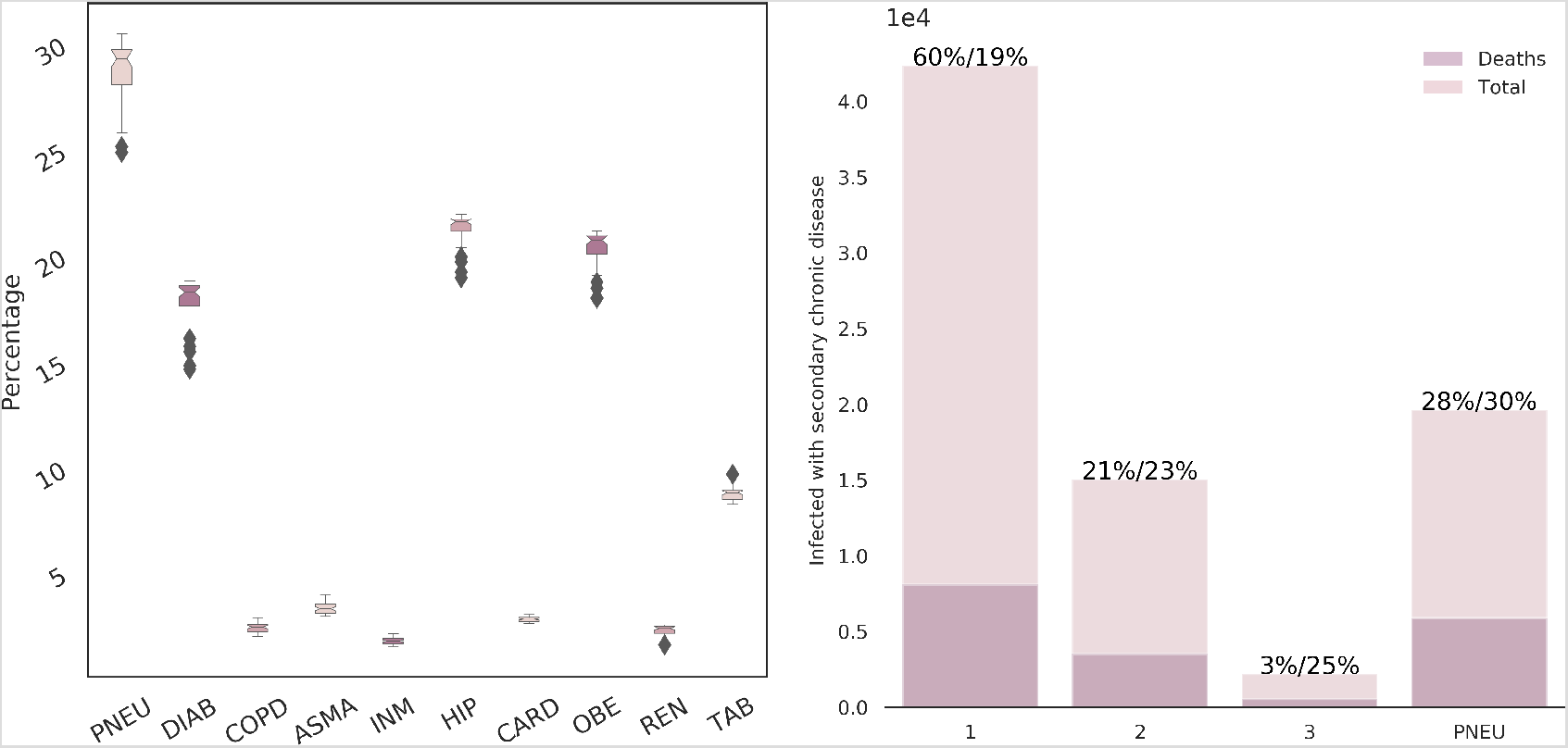
Reported data of most representative coexisting diseases. Data dispersion (left) shows the time percentage variability from April 12. Histogram in the right refers to the amount of individuals on May 25 having pneumonia (PNEU) or three of the chronic diseases with the higher incidence: diabetes, COPD and obesity. Labels 1, 2, and 3 represent the number of infected with one or the combination of 2 or 3 of the chronic diseases described above. Percentages on the top of the histogram columns are the percentage of the infected with the given condition referred to the total infected population and the mortality percentage for each category.

While keeping the elderly in social confinement until a vaccine is available, the best strategy to contain the epidemic in Mexico would be to return social activities after the first peak of the pandemic, returning the confined working class (about 27% of the population) in four equal parts, and at the last the returning of students to schools (see Figure 3(g)). Considering the best de-confinement strategy, Mexico would need at least a 3 fold increase in current hospital capacities. Note that this is an upper view of the pandemic at country level, however, the fate decision of pandemic evolution has shown great differences not only among countries but also among states inside of countries. Therefore, social confinement lifting should be tailored for each city in Mexico. Our results in Figure 4 highlight that Mexican Government may need to consider in de-confinement strategies not only on the age factor in COVID-19 patients but also people with chronic diseases such as diabetes, COPD and obesity. This is also a main concern, as in Mexico 65% of the population is overweight, 30% of the population is obese, and about 10% of the population had diabetes^25^.

In the same spirit of the epic phrase by Goerge E.P. Box that ”All models are wrong, but some are useful”, our model as well as others for COVID-19 has several limitations. In particular, mathematical models that are fitted during the emerging phase, exponential growth, of an outbreak that have potential sources of bias^26^. Furthermore, the data of infected cases in Mexico is very limited because of application of the sentinel model. While sampling is based on 10% of the suspected cases while 100% of hospitalized cases and deaths, mass testing for stage 2 and 3 is considered irrelevant. In fact, reports by the OECD (COVID-19)^27^ underlines that Mexico is only testing 0.6–0.8 per every thousand, occupying in this way Mexico the last place among countries with high number of infected cases. With 5,623 new tests on 4 May 2020, Mexico reported the highest number of samples per day^27^. Therefore, with this sample rate Mexico will be unable to register the dimension of the epidemic, actually, it is expected that Mexico will have a flat region as soon as the number of infected cases per day passes over 7,560. In reality, during the peak of the pandemic Mexico would need to test at least 18 per every thousand to provide a better vision of the problem.

With the progressing of information of COVID-19 pandemic as well as new data of infected cases in Mexico, our predictions would be more accurate and hopefully less drastic scenarios. Nevertheless, our predictions underline critical scenarios and suggest tailor public health strategies for social confinement lifting in combination with a significant increase in the health care system capacities.

## Data Availability

It is included.

## Author contributions statement

CLAR performed the simulations. EAHV envisaged the project . All the authors discussed and wrote the paper.

## Acknowledgements

This research was funded by the Universidad Nacional Autonoma de Mexico (UNAM), CONACYT, and the Alfons und Gertrud Kassel-Stiftung. This work received support from Luis Aguilar of the Laboratorio Nacional de Visualización Cientifica Avanzada (LAVIS) at UNAM-Juriquilla.

## References

1. CDC. Coronavirus diseases (COVID-2019) situation reports (2020).

2. Lowe, D. A Close Look at the Frontrunning Coronavirus Vaccines As of May 1 (updated) | In the Pipeline. Tech. Rep. (2020).

3. Anderson, R. M., Heesterbeek, H., Klinkenberg, D. & Hollingsworth, T. D. How will country-based mitigation measures influence the course of the COVID-19 epidemic? The Lancet DOI: 10.1016/S0140-6736(20)30567-5 (2020).

4. Kucharski, A. J. et al. Early dynamics of transmission and control of COVID-19: a mathematical modelling study. The Lancet Infect. Dis. 3099, 1–7, DOI: 10.1016/s1473-3099(20)30144-4 (2020).

5. Weitz, J. S. et al. Modeling shield immunity to reduce COVID-19 epidemic spread. Nat. Medicine DOI: 10.1038/s41591-020-0895-3 (2020).

6. Acuña-Zegarra, M. A., Santana-Cibrian, M. & Velasco-Hernandez, J. X. Modeling behavioral change and COVID-19 containment in Mexico: A trade-off between lockdown and compliance. Math. biosciences 108370, DOI: 10.1016/j.mbs.2020.108370 (2020).

7. Li, Q. et al. Early Transmission Dynamics in Wuhan, China, of Novel Coronavirus–Infected Pneumonia. New Engl. J. Medicine NEJMoa2001316, DOI: 10.1056/NEJMoa2001316 (2020).

8. Baud, D. et al. Real estimates of mortality following COVID-19 infection. The Lancet Infect. Dis. 3099, 30195, DOI: 10.1016/s1473-3099(20)30195-x (2020).

9. Li, R. et al. The demand for inpatient and ICU beds for COVID-19 in the US: lessons from Chinese cities. medRxiv 2020.03.09.20033241, DOI: 10.1101/2020.03.09.20033241 (2020).

10. Kissler, S. M., Tedijanto, C., Goldstein, E., Grad, Y. H. & Lipsitch, M. Projecting the transmission dynamics of SARS-CoV-2 through the postpandemic period. Science DOI: 10.1126/SCIENCE.ABB5793 (2020).

11. INEGI. Encuesta Nacional de Ocupación y Empleo (ENOE), población de 15 años y más de edad. Tech. Rep. (2019).

12. Secretaria de Salud MEXICO. Coronavirus (COVID-19)-Comunicado Técnico Diario | Secretaría de Salud | Gobierno | gob.mx. Tech. Rep. (2020).

13. Secretaría de Salud. Manual para la vigilancia epidemiológica de Influenza. Dirección Gen. de Epidemiol. 1–84 (2014).

14. Peng, L., Yang, W., Zhang, D., Zhuge, C. & Hong, L. Epidemic analysis of COVID-19 in China by dynamical modeling. arXiv (2020). 2002.06563.

15. Verity, R. et al. Estimates of the severity of coronavirus disease 2019 : a model-based analysis. Lancet Infect. Dis. 3099, 1–9, DOI: 10.1016/S1473-3099(20)30243-7 (2020).

16. Wölfel, R. et al. Virological assessment of hospitalized patients with COVID-2019. Nature 1–10, DOI: 10.1038/s41586-020-2196-x (2020).

17. WHO. WHO Director-General’s opening remarks at the media briefing on COVID-19 – 24 February 2020 (2020).

18. Phua, J. et al. Intensive care management of coronavirus disease 2019 (COVID-19): challenges and recommendations. The Lancet Respir. Medicine 8, 506–517, DOI: 10.1016/s2213-2600(20)30161-2 (2020).

19. Reporte de la Secretaria de Salud MEXICO (8 de Abril 2020). Versión estenográfica. Conferencia de prensa. Informe diario sobre coronavirus COVID-19 en México (2020).

20. Google. COVID-19 Community Mobility Reports.

21. Storn, R. & Price, K. Differential Evolution – A simple and efficient adaptive scheme for global optimization over continuous spaces. J. Glob. Optim. 11, 341–359, DOI: 10.1023/A:1008202821328 (1997).

22. Parra-Rojas, C. & Hernandez-Vargas, E. A. PDEparams: parameter fitting toolbox for partial differential equations in python. Bioinformatics DOI: 10.1093/bioinformatics/btz938 (2019).

23. Dirección General de Epidemiología. Datos Abiertos para COVID-19 – Dirección General de Epidemiología | Secretaría de Salud | Gobierno | gob.mx.

24. Velázquez Ramírez, M. d. C. Capacidad instalada en México para enfrentar al coronavirus COVID-19 (2020).

25. Levaillant, M., Lièvre, G. & Baert, G. Ending diabetes in Mexico. Lancet (London, England) 394, 467–468, DOI: 10.1016/S0140-6736(19)31662-9 (2019).

26. Britton, T. & Tomba, G. S. Estimation in emerging epidemics: biases and remedies. J. Royal Soc. Interface 16, DOI: 10.1098/RSIF.2018.0670 (2019).

27. OECD. Testing for COVID-19 : A way to lift confinement restrictions. Tech. Rep. April, OECD (2020).

